# The Impact of Brain-Derived Neurotrophic Factor Polymorphism and Stimulation Parameters on the Response to Repetitive Transcranial Magnetic Stimulation: A Systematic Review

**DOI:** 10.1101/2024.11.06.24316617

**Authors:** Yi-Ling Kuo, Gracy Lin, Stephen J. Glatt

## Abstract

**Introduction:** TMS has been a common technique used to stimulate neuromodulatory changes, which can have therapeutic effects. The underlying mechanism is still unknown, however it is thought to cause neuroplastic changes via LTD or LTP. However, the effects are highly variable, with demographics and baseline physiology thought to be playing a role.

**Objectives:** The purposes of this systematic review were to 1) examine how BDNF polymorphisms are related to the after-effects of rTMS in humans and 2) investigate the association between BDNF polymorphism and rTMS stimulation parameters as contributing factors to the response to rTMS.

**Materials and Methods:** Studies identified from PubMed, The Cochrane Library, and Embase were screened for eligibility. Data were extracted from the selected studies by one reviewer and verified by another reviewer. Risk of Bias was assessed using the Cochrane Collaboration’s tool. Results were synthesized narratively.

**Results:** Of the 224 initial studies, 35 were included in this systematic review. 33 out of 35 studies had at least one domain of high or unclear risk of bias. 53% of the studies in healthy individuals showed differences in TMS-derived or behavioral measures between Val/Val homozygotes and Met allele carriers. The neuromodulatory effects were more reliable in Val/Val homozygotes than Met allele carriers. In stroke, neuromodulatory effects on corticospinal excitability and motor deficits were more evident in Val/Val homozygotes than Met allele carriers. Similarly, in depression, Val/Val homozygotes demonstrated more improvement in depression symptoms compared with Met allele carriers following rTMS. The role of BDNF polymorphism in other disorders remained unclear.

**Conclusion:** It remains inconclusive whether and how BDNF genotype impacts the effects of rTMS. Methodological heterogeneity in the stimulation parameters, such as dosage and excitatory or inhibitory protocols, interact with BDNF polymorphism and contribute to the response to rTMS.

## INTRODUCTION

The rapid increase in the application of non-invasive brain stimulation (NIBS) has led to numerous breakthroughs in experimental and clinical neuroscience. Transcranial magnetic stimulation (TMS) has been the most used NIBS technique for studying cortical physiology and neuroplasticity. A high-current generator flows electrical current to the conducting coil and generates a magnetic field that induces an electric current to depolarize superficial axons and create action potentials in the cortex^1,2^. The repetitive introduction of TMS pulses can alter brain function lasting for a period of time following the stimulation, typically tens of minutes, which is known as repetitive TMS (rTMS). The neuromodulatory effects of rTMS are dependent on the stimulation site, frequency, intensity of the magnetic pulses, and orientation of the coil. The consensus from the neuromodulation societies considers low-frequency stimulation of ≤1 Hz to be “inhibitory’’, whereas high-frequency stimulation of ≥5 Hz to be ‘‘excitatory” in modulating corticospinal excitability (CSE). The modified form of rTMS with a much higher stimulation frequency, called theta-burst stimulation (TBS), produces a longer-lasting neuromodulatory effect (up to 60 minutes) within a shorter stimulation time frame and at a lower intensity^3^. TBS is administered as a continuous (cTBS) train with inhibitory effects, or intermittent (iTBS) train with excitatory effects^1^. It is generally acknowledged that TBS modifies CSE with a robust change similar to the cellular processing of long-term depression (LTD) with an inhibitory effect and long-term potentiation (LTP) with an excitatory effect^1,4^.

The mechanism underlying the neuroplastic changes associated with the transient or lasting rTMS- modulated CSE remains unclear^4^. rTMS-induced neuromodulatory effects are highly variable and it has been demonstrated that only approximately 50% of participants responded to TBS with anticipated experimental and therapeutic effects^5^. Inter- and intra-individual variability of rTMS is dependent on baseline demographics and physiological characteristics such as anthropometric measurements of the head and brain, pathological changes in the central and peripheral nervous systems, and past life experiences leading to neuroplastic changes^6^. rTMS is used as a treatment to intervene in the brain circuits and has been shown to demonstrate efficacy in psychiatric and neurologic disorders^7^. Even though rTMS has been FDA-approved for a few indications, lacking markers to predict treatment efficacy is a long-standing issue in the field of neuromodulation. It is important to identify potential responders to rTMS to enhance the cost-effectiveness of this technique given the highly variable nature of this technique^4^. Predictor studies will help to pinpoint individuals suitable for rTMS intervention. Demographics, neurophysiology, and genetic traits have been studied. Using demographic, neurobiological, and clinical factors as predictors of rTMS treatment response is critical to fill the knowledge gap in identifying candidates to receive rTMS treatment.

Brain-derived neurotrophic factor (*BDNF*) is one of the candidate genes that contribute to brain morphological and functional changes and plays a critical role in synaptic plasticity and neuroprotection, integrating the metabolic and behavioral responses to the environments^8,9^. The Val66Met (rs6265) variant is a functional nonsynonymous single nucleotide polymorphism (SNP) in the *BDNF* gene that encodes the *BDNF* protein on chromosome 11 (11p13). This SNP of *BDNF* is hypothesized to affect the heterogeneity of response to rTMS as the placement of Val or Met occurring at position 66 in the pro-region of the *BDNF* gene determines the distribution of *BDNF* in the brain^10^. As *BDNF* is essential in regulating synaptic plasticity and brain connectivity, the Val66Met polymorphism is associated with reduced activity-dependent *BDNF* release, which can lead to cognitive deficits and susceptibility to neurologic and psychiatric disorders^11,12^. Another frequently studied *BDNF* polymorphism is C270T (rs56164415), a variant of a C to T substitution in the 5D untranslated region that impacts the transcription of the *BDNF* gene. *BDNF* C270T alters the efficacy of *BDNF* translation, which can lead to *BDNF* imbalance in the brain and has also been linked to neurologic and psychiatric disorders^13^.

There are increasing studies suggesting that *BDNF* polymorphism is a vital factor in identifying responders to rTMS. However, there is conflicting evidence on the impact of *BDNF* genotype on responsiveness to rTMS, which might be attributed to various physiological factors and stimulation parameters. Here, this systematic review aims to 1) examine how the rs6265 and rs56164415 *BDNF* polymorphisms are related to the after-effects of rTMS in humans and 2) investigate the association between *BDNF* polymorphism and rTMS stimulation parameters as contributing factors to the response to rTMS.

## MATERIALS AND METHODS

### 2.1 Protocol

The guidelines of the Preferred Reporting Items for Systematic Reviews and Meta-analyses for Protocols 2015 (PRISMAm-P 2015) were used to conduct this systematic review. The protocol was registered in the PROSPERO.

### 2.2 Eligibility Criteria

Studies met the following eligibility criteria were included in this systematic review: (1) research in humans; (2) peer-reviewed studies published in English; (3) studies with accessible full-text; (4) original research; (5) participants classified and grouped according to *BDNF* genotype; (6) rTMS used as the non-invasive brain stimulation intervention; (7) outcomes measured pre and post rTMS intervention to determine changes and treatment effects; (8) outcomes compared between groups with different *BDNF* genotypes or between groups receiving active and sham rTMS.

### 2.3 Search Strategy

Electronic literature searches were performed using PubMed, The Cochrane Library, and Embase, up to November 2024. These databases were searched using keywords and medical subject headings (MeSH), including: ‘rTMS’ or ‘repetitive transcranial magnetic stimulation’ or ‘TBS’ or ‘theta burst stimulation’ and ‘*BDNF*’ or ‘brain-derived neurotrophic factor’ and ‘polymorphism’ or ‘genotype’. Titles and abstracts of the initial search results were screened for relevant studies meeting the inclusion criteria. Full-text versions of potentially eligible studies were then examined. A manual search of the reference lists of the eligible studies was also performed.

### 2.4 Selection Process

One reviewer (YLK) initially screened the titles and abstracts of the articles, and the eligibility of the screened studies was determined independently by all reviewers.

### 2.5 Outcome Measures

The effects of rTMS or TBS on TMS-measured motor evoked potential (MEP) amplitudes were included as the neurophysiological outcomes. Outcomes other than TMS-derived measurements were included as behavioral outcomes or clinical outcomes.

### 2.6 Data Extraction

Demographic data extracted included the study population, sample size of each group, mean age, and sex. Brain stimulation-related data included the location of stimulation, side of stimulation (left or right hemisphere), stimulation protocol (stimulation frequency, session number, stimulation intensity, and number of stimuli), and sham stimulation. Methods of obtaining body fluid drawing and *BDNF* genotyping were extracted. Behavioral and clinical outcome measures obtained before and after rTMS or TBS were extracted. Data were independently extracted by one reviewer and verified by another reviewer (YLK and GL). The reviewers discussed any discrepancy in data to reach a consensus.

### 2.7 Risk of Bias

The Cochrane Collaboration’s tool^14^ was used to assess the risk of bias in each included study. The domains assessed the risk of selection bias, performance bias, detection bias, attrition bias, reporting bias, and other biases. The ratings were categorized as low, unclear, or high risk of bias. The risk of bias in each included study was independently assessed by one reviewer and verified by another reviewer (GL and YLK).

## RESULTS

### 3.1 Selection of Studies

Electronic literature searches with keyword searches yielded a total of 224 studies. After removing the duplicates, 115 studies were included in the initial screening of titles and abstracts with the eligibility criteria. 74 studies were eliminated due to the following reasons: (1) studies were conference abstracts or proceedings (N = 16); (2) studies were reviews or commentaries (N = 18); (3) rTMS or TBS was not used as an intervention (N = 32); (4) articles were not written in English (N = 2); (5) records were protocol registrations (N = 6); (6) studies were performed in animals (N = 5); and (7) participants all had the same *BDNF* genotype and all received iTBS without a control condition (N = 1). As a result, 35 articles met the full eligibility criteria and were included in this systematic review. The selected studies were categorized according to population, including 17 studies in healthy individuals, nine in stroke, three in depression, one in spinocerebellar ataxia, one in Parkinson’s Disease with levodopa-induced dyskinesias, one in Tourette syndrome, one in schizophrenia, one in autism spectrum disorder, and one with combined populations of older adults, Type-2 diabetes mellitus, and Alzheimer’s disease. The PRISMA diagram (Figure 1) demonstrates the study inclusion process.

**Figure 1.**
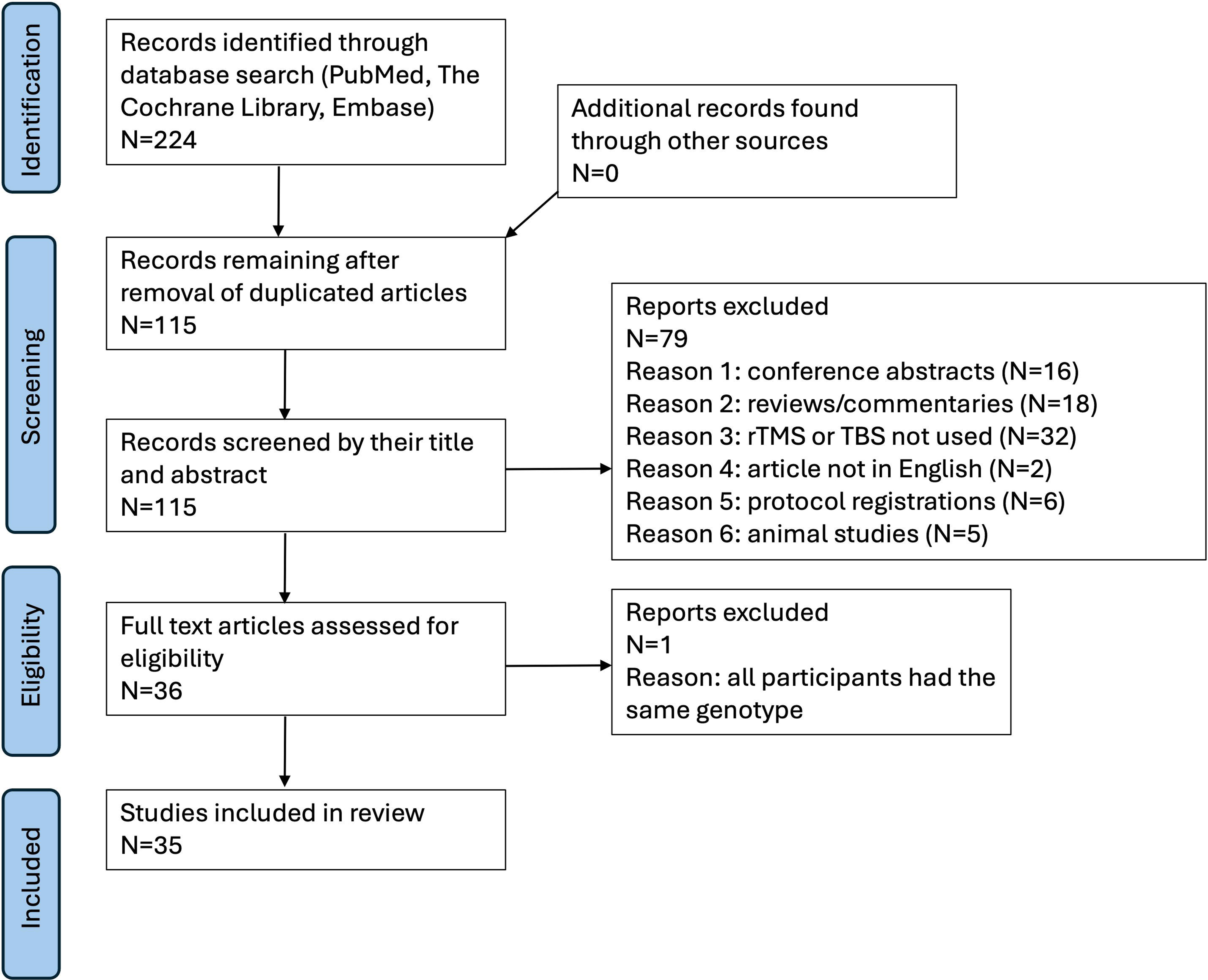
PRISMA diagram of the review

### 3.2 Risk of Bias

The risk of bias in the studies grouped by participant condition is presented in Figure 2. Risk of bias categories were “low risk of bias,” “unclear risk of bias,” or “high risk of bias.” Most studies had a high or unclear risk of bias in three or more domains. Out of the total 35 studies, two had a high risk of bias for not using random sequence, five had a high risk of bias for not using allocation concealment, two had a high risk of bias for not blinding participants and researchers, one had a high risk of bias for not blinding outcomes assessment, 16 had a high risk of bias for incomplete outcome data, one had a high risk of selective reporting, and 17 had a high risk of other bias. Most studies did not include information regarding allocation concealment and blinding of outcome assessment. 33 out of 35 (94.3%) studies had at least one domain of unclear risk of bias.

**Figure 2.**
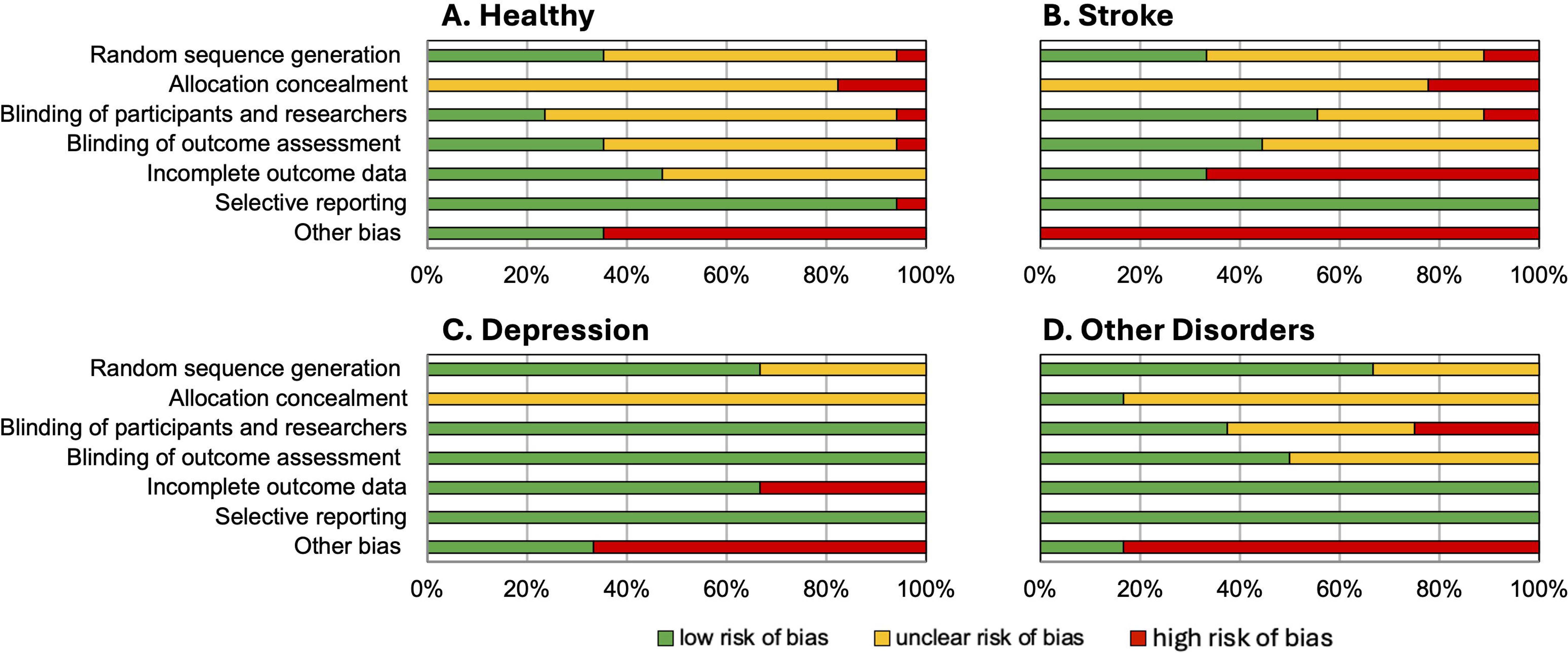
Risk of bias assessments showing the percentage of included studies with high, unclear, or low risk of bias in the population of A. healthy individuals; B. stroke; C. depression; and D. other disorders.

### 3.3 *BDNF* Genotyping Methods

Twelve studies obtained saliva samples to perform *BDNF* genotyping^15–26^, whereas other studies used venous blood samples. *BDNF* Val66Met polymorphisms were categorized as homozygous for valine in codon 66 (Val/Val), homozygous for methionine (Met), or Val/Met heterozygotes. Met/Met homozygotes were less seen in the general population, and only four studies had sufficient sample size to group and compare participants across all polymorphisms (Val/Val versus Val/Met versus Met/Met)^27–30^. Individuals with variants of Val/Met and Met/Met were pooled as Met allele carriers in some studies to refer to individuals with the Met variant form of *BDNF*^16,20,25,31–40^. Only two studies genotyped the C270T polymorphisms: one in stroke^34^ and one in schizophrenia^41^.

### 3.4 Stimulation Parameters, *BDNF* Polymorphism, and Responses to rTMS

Data extracted from the included studies are presented in Table 1. Low-frequency rTMS studies used 1 Hz, whereas high-frequency rTMS studies used 5, 10, and 20 Hz. TBS studies used 50 Hz as the stimulation frequency in both iTBS and cTBS. The stimulation sites included M1, frontal cortex, dorsal lateral prefrontal cortex (DLPFC), Broca’s area, and cerebellum, with M1 being the most stimulated location. For single-session studies examining the immediate effects of rTMS and TBS, stimulation stimuli applied ranged from 250 to 1,500. For multi-session studies, rTMS and TBS were used as therapeutic interventions with neuromodulatory effects in some studies on stroke^30,34,35,37,42^ and depression^39,43,44^, as well as other disorders^26,41,45^. The treatment courses were five to 22 sessions over one to three weeks, with stimulation stimuli ranging from 300 to 1,800 per session.

**Table 1.**
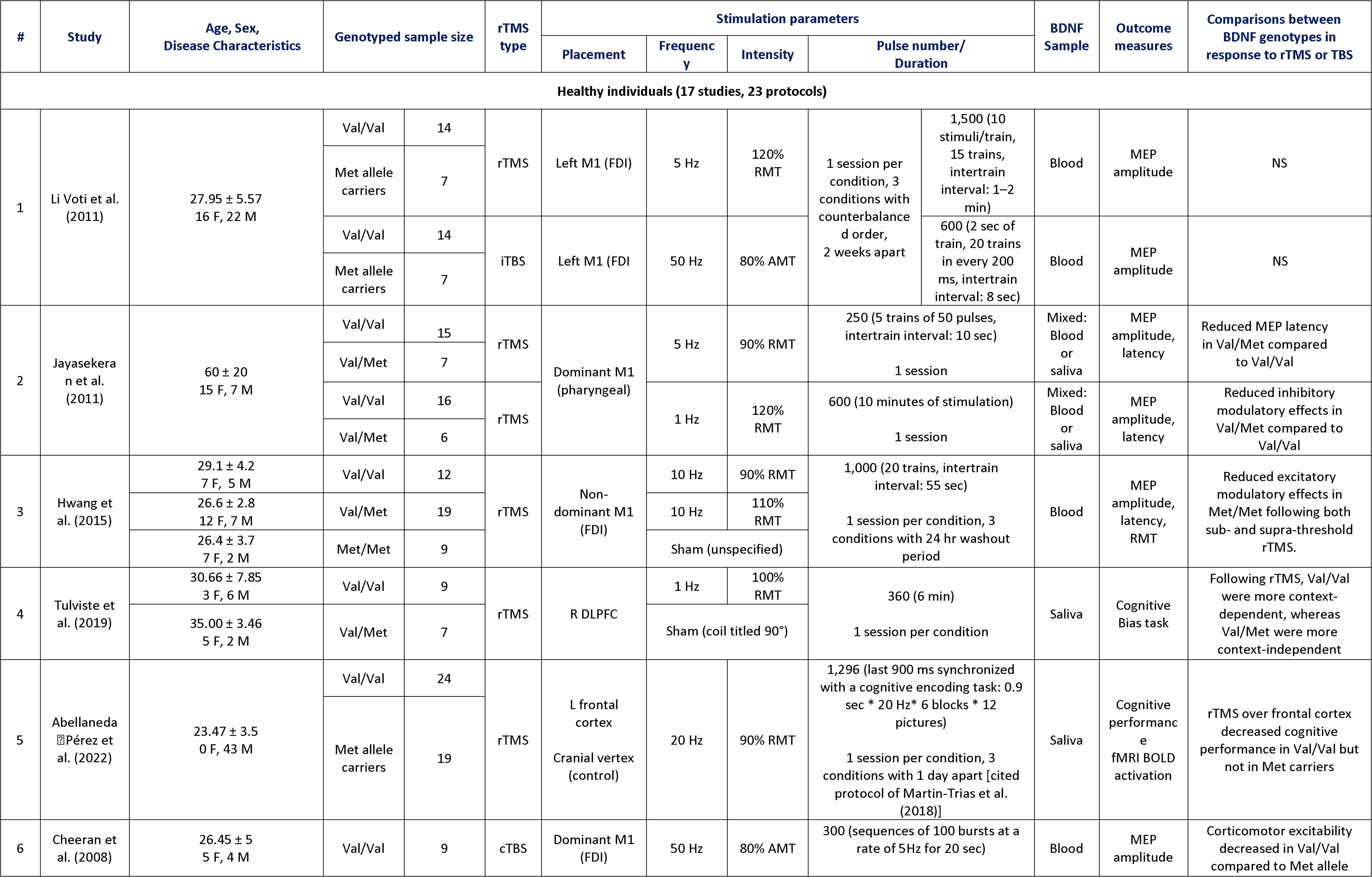
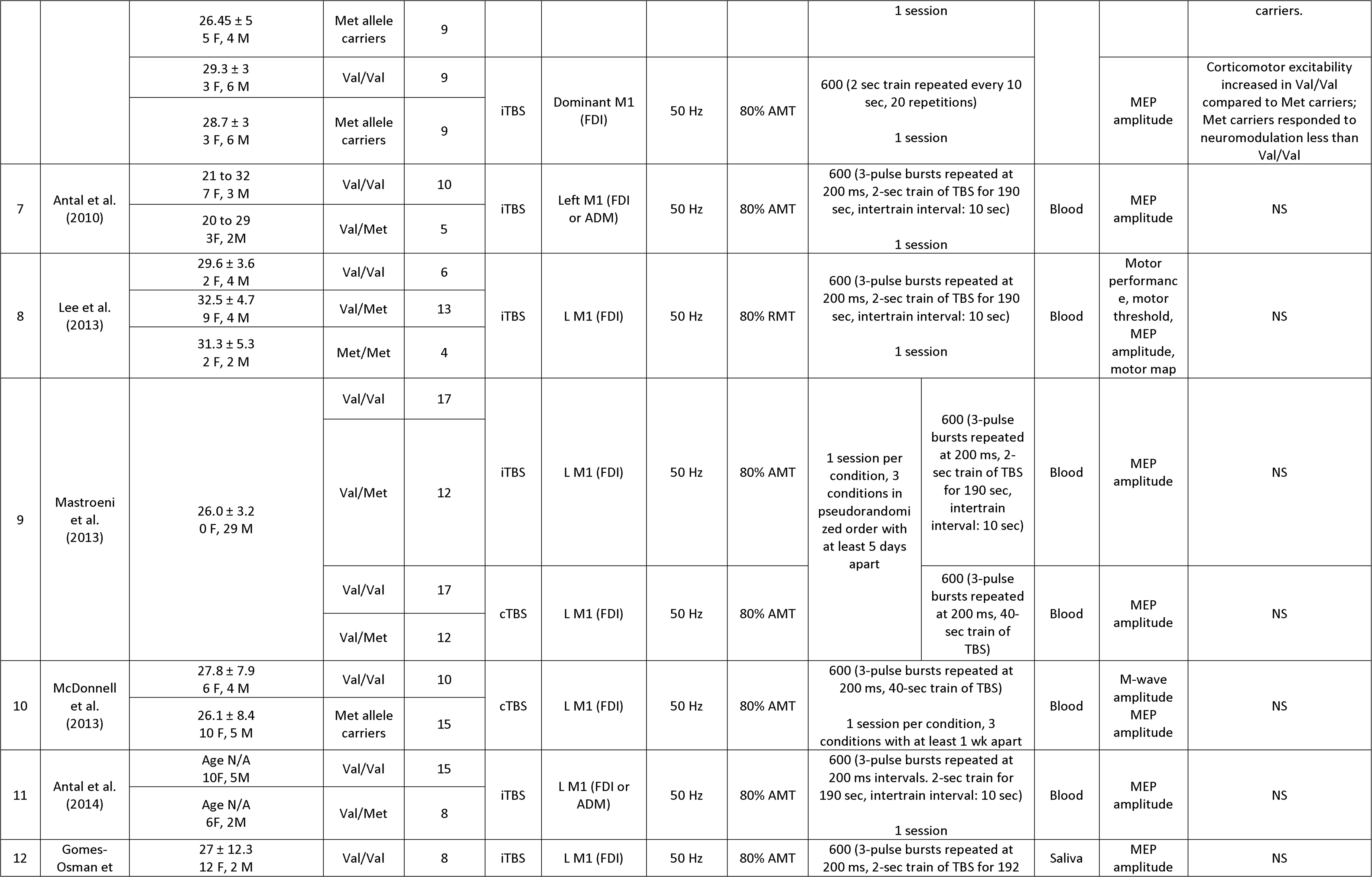
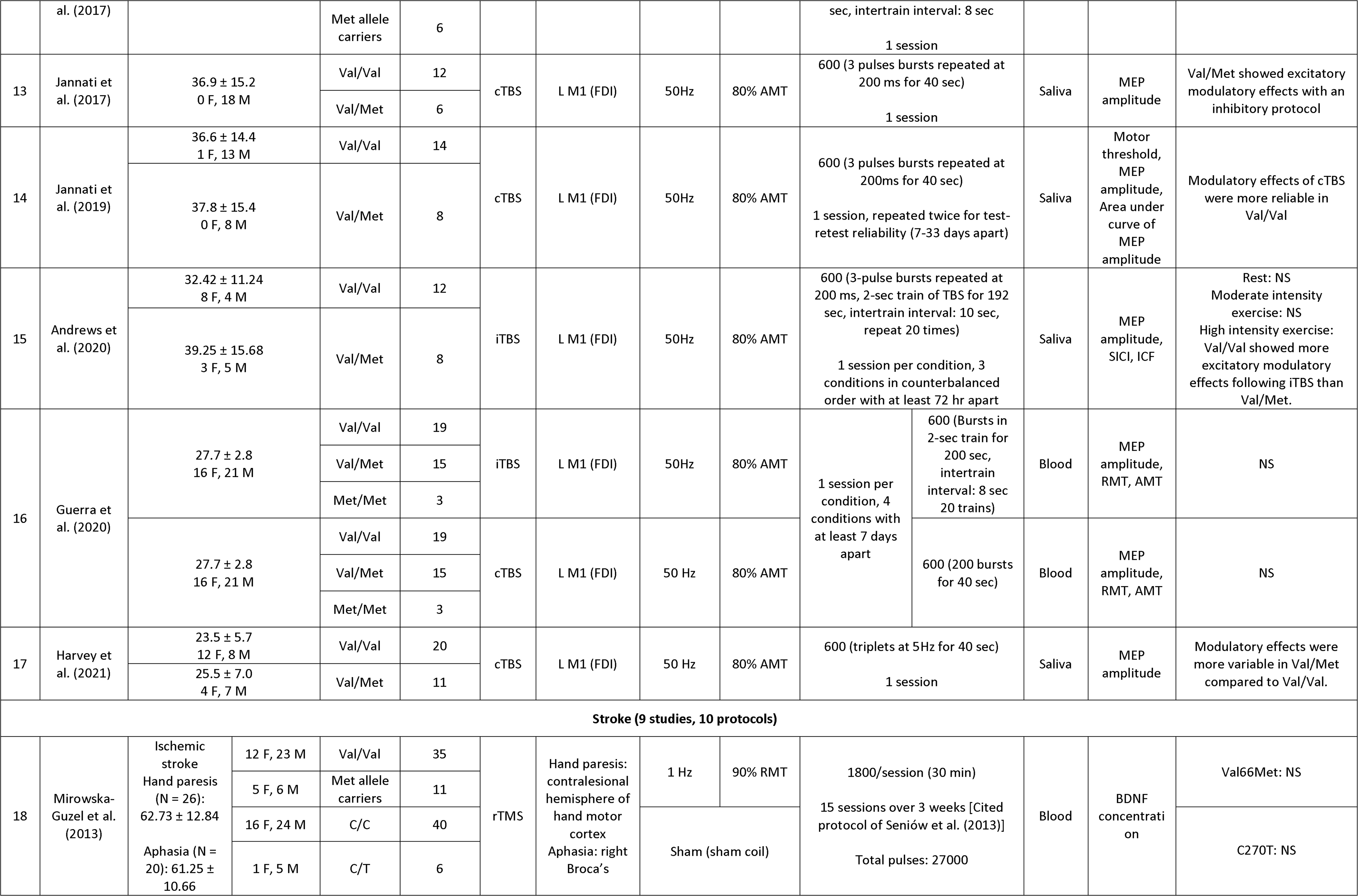
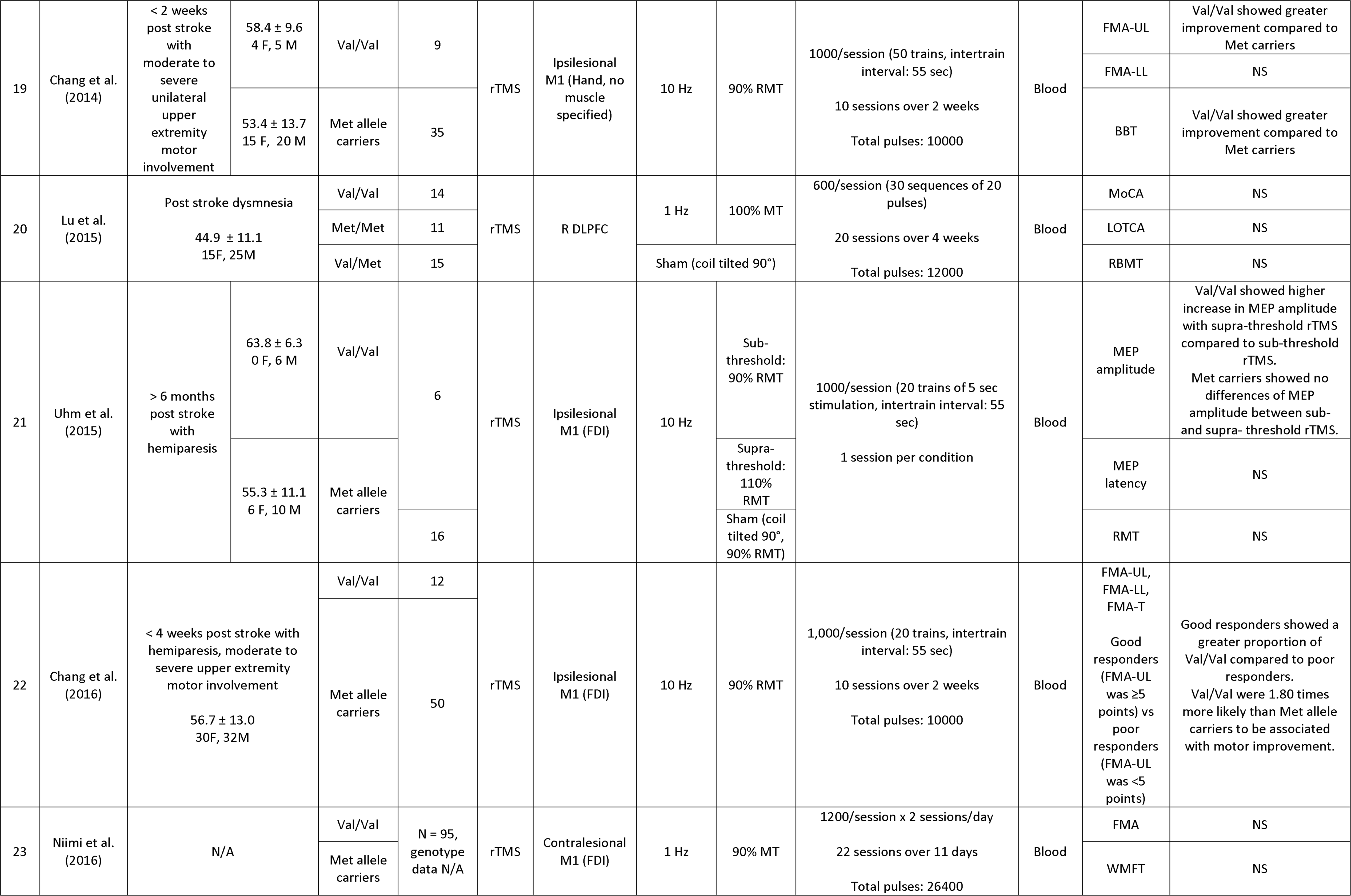
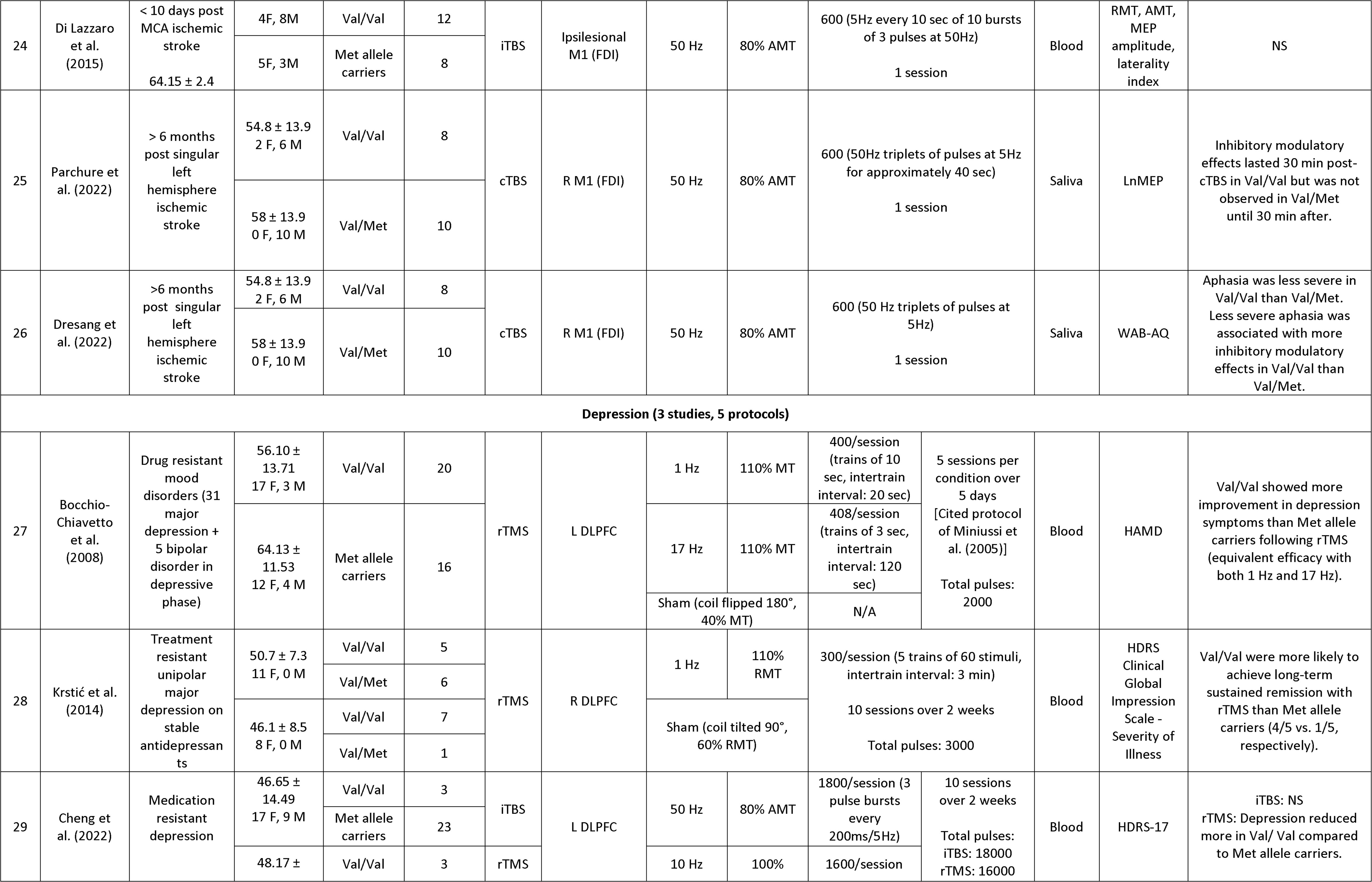
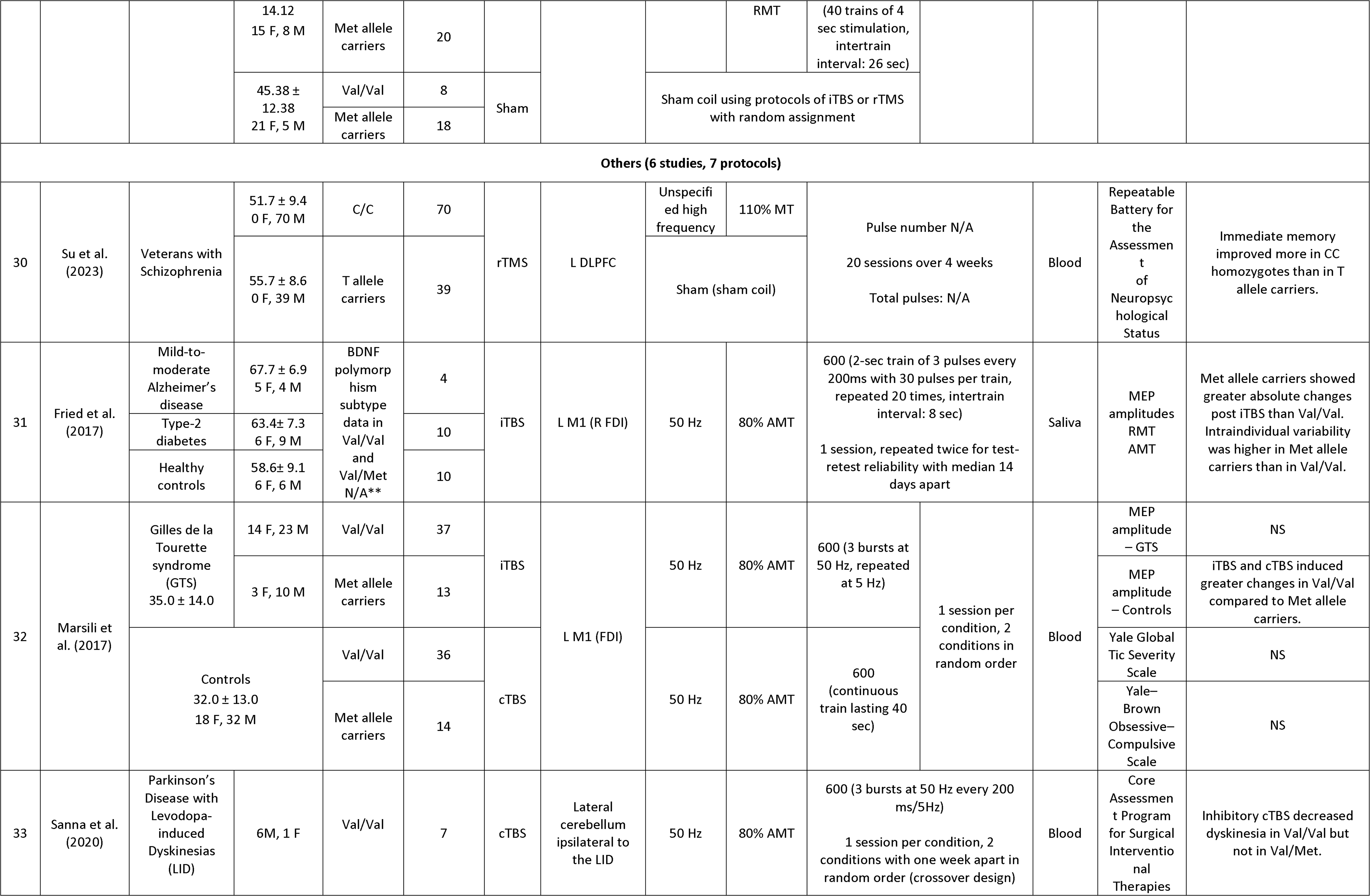
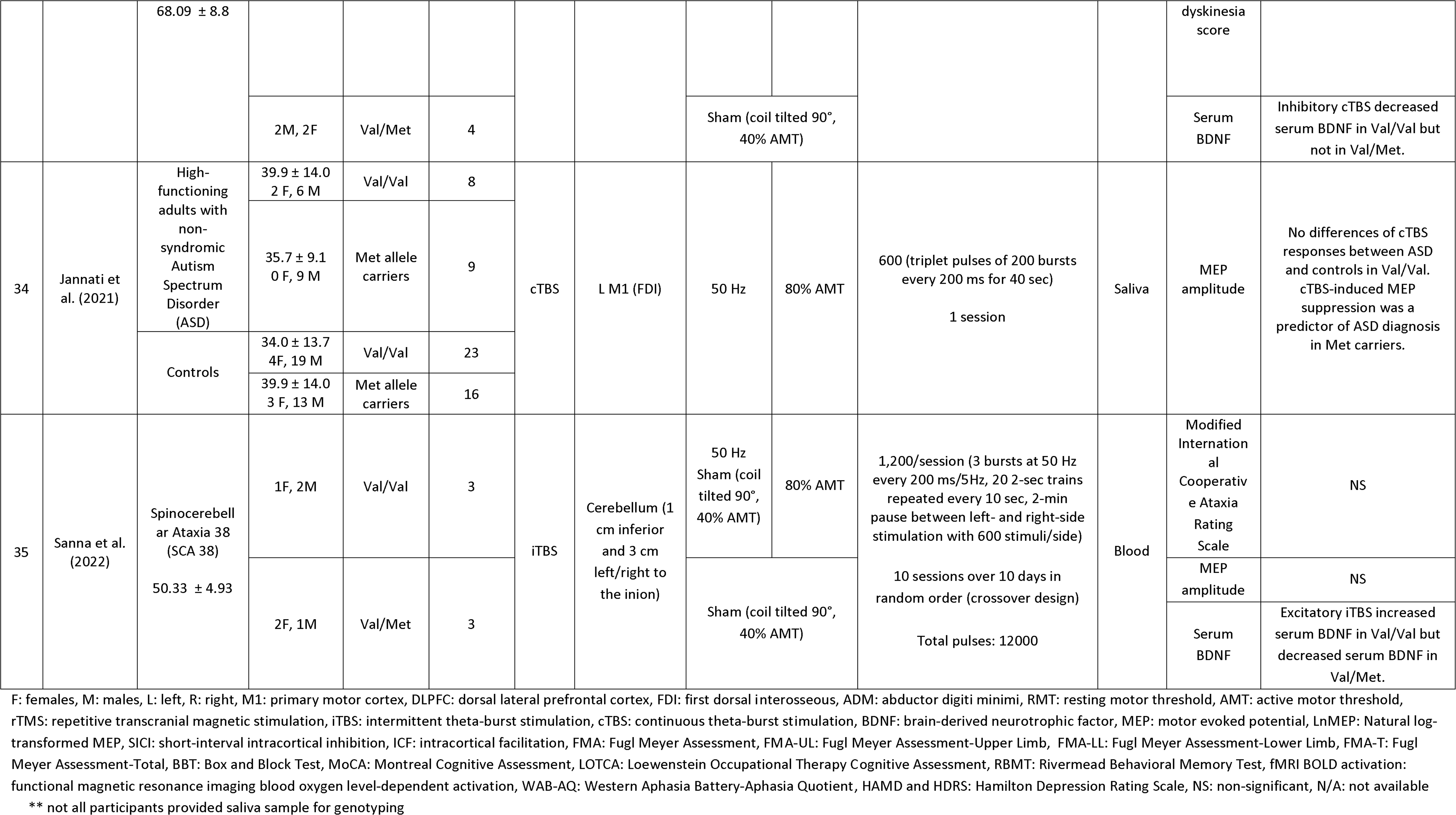
Data extraction.

#### Healthy Individuals

Most studies in healthy individuals targeted the left M1 at the representational area of the first dorsal interosseus (FDI) muscle, whereas some studies targeted the dominant M1 of the pharyngeal muscle^17^, dominant^32^ or non-dominant^27^ M1 of the FDI, right DLPFC^15^, and left frontal cortex^16^. Five studies administered rTMS (high and/or low frequency)^15–17,27,31^, and 14 studies administered TBS (iTBS and/or cTBS)^18–22,28,29,31–33,46–49^. Sub-threshold^16,17,27,34–37,42^, at-threshold^15,30,39^, or supra- threshold^17,27,30,31,36,39,41,43,44^ intensities were used in rTMS protocols, ranging from 90% to 120% resting motor threshold (RMT) or motor threshold (MT) without specifying muscle activation status. TBS protocols, including iTBS and cTBS, used an 80% active motor threshold (AMT) as the stimulus intensity, except for one study using 80% RMT^28^.

Most studies used a single-session cross-sectional design to investigate the acute effects of rTMS or TBS. MEP amplitude was the most commonly used outcome measure to capture brain physiology^17–22,27–29,31–33,46,47,49^, while some studies reported the neuromodulatory effects with other physiological parameters such as MEP latency^17,27^, RMT^27–29^, AMT^29^, short-interval intracortical inhibition^18^, intracortical facilitation^18^, and motor map^28^. Behavior outcomes measuring cognitive function were used in two rTMS studies^15,16^. Some studies showed that neuromodulatory effects were less observed in Met allele carriers compared to Val/Val homozygotes^17,18,22,27,32^. Val/Val homozygotes demonstrated more reliable neuromodulatory effects (excitatory or inhibitory) than Met allele carriers^21,22,26^. Alterations in behaviors measured by cognitive performance were evident in Val/Val homozygotes but not in Met allele carriers^15,16^. However, 47% of the studies in healthy individuals did not observe differences in neuromodulatory effects between Val/Val homozygotes and Met allele carriers, of which one study used rTMS^31^, seven studies used iTBS^20,28,29,31,46,47,49^, and three studies used cTBS^29,33,46^.

#### Stroke

In stroke, three studies used high-frequency (10 Hz) rTMS at the ipsilesional M1^35,36,42^; two used low-frequency rTMS at the contralesional M1^34,37^; one used iTBS at the ipsilesional M1^38^; and two used cTBS at the right M1^23,24^. Only one study targeted the right DLPFC with low-frequency rTMS^30^. Neuromodulatory effects on CSE were greater in Val/Val homozygotes compared to Met allele carriers^23,36^. Motor impairments of the upper limb measured by the Fugl Meyer Assessment and Box and Block Test were improved with excitatory high-frequency rTMS applied to the ipsilesional M1 in Val/Val homozygotes compared to Met allele carriers^35,42^, which was not observed with inhibitory low-frequency rTMS applied to the contralesional M1^37^. Inhibitory cTBS reduced aphasia more in Val/Val homozygotes than Val/Met heterozygotes^24^. However, *BDNF* polymorphism has no impact on the CSE, *BDNF* concentration, and cognitive symptoms following low-frequency rTMS^30,34^ or iTBS^38^ in stroke.

#### Depression

All three studies on depression stimulated the DLPFC, and two studies targeted the left^39,43^ while one targeted the right DLPFC^44^. Low-frequency rTMS^43,44^, high-frequency rTMS^39,43^, and iTBS^39^ were used in these studies on depression. Stimulation intensities ranged from 100% to 110% RMT in rTMS studies^39,43,44^. A total of five to 10 sessions were administered with the number of stimulation stimuli ranging from 300 to 1800 per session. All rTMS studies showed more improvement in depression symptoms (measured by the Hamilton Depression Rating Scale) in Val/Val homozygotes than Met allele carriers^39,43,44^, whereas *BDNF* polymorphisms did not affect the change of depression symptoms following iTBS^39^.

#### Other Disorders

In studies on disorders other than stroke or depression, iTBS or cTBS was used on the left M1 of the FDI^25,26,40^. Two studies used iTBS^45^ or cTBS^50^ targeting the cerebellum, and one used high-frequency rTMS on the left DLPFC^41^. There were various outcome measures used to capture the neuromodulatory effects of rTMS and TBS, including physiological measures such as MEP amplitudes and RMT, and behavioral or clinical measures of symptoms. One study in Parkinson’s Disease with levodopa-induced dyskinesias^50^ and another study in Spinocerebellar Ataxia 38^45^ demonstrated greater neuromodulatory responses on serum *BDNF* with Val/Val homozygotes compared to Val/Met heterozygotes. Nonetheless, studies on Gilles de la Tourette syndrome and Spinocerebellar Ataxia 38 did not find different responses to TBS in clinical symptoms and MEP amplitudes between Val/Val homozygotes and Met allele carriers^40,45^. In this systematic review, only one study demonstrated greater neuromodulatory effects in Met allele carriers than Val/Val homozygotes after pooling the data from Alzheimer’s disease, Type-2 diabetes mellitus, and controls^26^. Val/Met heterozygotes may result in more variability in the response to neuromodulation, which is consistent with the finding of another study in healthy individuals^21^.

## DISCUSSION

This systematic review investigated the impact of *BDNF* genotypes, including the rs6265 and rs56164415, on the response to rTMS or TBS in the literature of human research. There was high heterogeneity in the research methodology across studies. 12 out of 23 (52.2%) of the rTMS or TBS protocols in healthy individuals demonstrated greater neuromodulatory effects with rTMS or TBS on neurophysiological measures such as MEP amplitude and behavioral measures of cognitive performance, in Val/Val homozygotes than Met allele carriers. 3 out of 10 (30.0%) protocols in stroke showed greater neuromodulatory effects on physiological measures like MEP amplitude or peripheral *BDNF* serum concentration, and another 30% of protocols showed more improvement in clinical outcomes with symptom improvement in Val/Val homozygotes compared to Met allele carriers. 4 out of 5 protocols in depression demonstrated more improvement in clinical outcomes with symptom improvement in Val/Val homozygotes compared to Met allele carriers. 11 out of 23 (47.8%) rTMS or TBS protocols in healthy individuals and 4 out of 10 (40.0%) protocols in stroke did not demonstrate different responses to rTMS or TBS in any outcome measures with the Val66Met polymorphism. The impact of *BDNF* Val66Met polymorphisms on other disorders was mixed.

### 4.1 Dosage

Determining appropriate dosage in the use of neuromodulation has been challenging due to a lack of standard definition of dosage^51^. The total number of pulses is the most common method of determining the dosage of TMS, while other parameters, such as stimulation intensity, stimulation frequency, and trial session number, contribute to the dose-response algorithm^51–53^. To date, the interaction between *BDNF* polymorphism and dosage of TMS has not been systematically investigated. Dosage can be discussed in two categories in the studies included in this systematic review: 1) immediate effect with a single session of neuromodulation and 2) therapeutic effect with multiple neuromodulation sessions. All studies in healthy individuals were designed to investigate the immediate effect of rTMS/TBS with a single session, and the number of stimuli ranged from 250 to 1500. Five protocols for stroke and five protocols for other disorders used a single-session design, with the number of stimuli ranging from 600 to 1000. Single-session rTMS/TBS has not been shown to consistently modulate CSE in healthy individuals^54^. Similarly, the current systematic review also showed that the number of stimuli did not appear to contribute to the differences observed between *BDNF* genotypes with the immediate after- effects of rTMS/TBS.

For multiple-session studies, five protocols for stroke, five protocols for depression, and two protocols for other disorders used five to 22 sessions to build up therapeutic effects with rTMS/TBS. There was high variability in the total number of stimuli used across these studies, between 2000 and 27000. Surprisingly, a lower total number of stimuli (10000 or less) appeared to be associated with differences observed in MEP amplitude and clinical outcomes between *BDNF* genotypes^35,42–44^, whereas a larger total number of stimuli (more than 10000) was associated with insignificant between-genotype differences^30,34,37,39,45^. It is possible that the influence of the *BDNF* polymorphism can be overcome by increasing the dosage with a large total number of stimuli to achieve the modulatory effects. However, *BDNF* polymorphism effects were not attenuated by higher dosages from increased stimulation intensity (100% RMT or higher)^15,17,27,36,39,41,43,44^.

### 4.2 Forms of rTMS

Theta burst stimulation, a modified form of rTMS, generates more substantial neuromodulatory effects in modulating CSE and synaptic plasticity than conventional rTMS^2,55^. It has been shown that the iTBS leads to longer-lasting increased CSE than high-frequency rTMS. Similarly, cTBS leads to longer-lasting decreased CSE than low-frequency rTMS, although the effects are less consistent than iTBS^2^. A previous systematic review and meta-analysis examined how *BDNF* polymorphism influenced TBS effects and demonstrated larger effect sizes in Val/Val homozygotes. Met allele carriers, on the other hand, demonstrated variable results in response to TBS^53^. Two studies included in the current systematic review demonstrated variable findings in Met allele carriers and also used TBS^21,26^. In studies with both rTMS and TBS protocols, one study in healthy individuals did not find *BDNF* polymorphism to impact CSE with either protocol^31^, whereas another study in depression showed that the depression symptoms reduced in Val/Val homozygotes but not Met allele carriers only with rTMS^39^. Overall, *BDNF* polymorphism influences response to neuromodulation in 14 out of 19 studies in rTMS and 14 out of 26 studies in TBS. It is possible that the impact of the *BDNF* polymorphism could be attenuated by a modified yet intensive form of rTMS.

### 4.3 Excitatory versus Inhibitory Protocols

There were conflicting findings on the consistency in modulating CSE with the excitatory versus inhibitory protocols in rTMS^56^ and TBS^53,57^. There are known demographic and personal factors contributing to the neuromodulatory effects of long-term depression and long-term potentiation, including *BDNF* polymorphism^9,58^. In the current systematic review, the *BDNF* polymorphism effect was evident in similar percentages of studies in excitatory and inhibitory protocols. Overall, 16 out of 26 (61.5%) excitatory protocols (including high-frequency rTMS and iTBS) and 13 out of 19 (68.4%) inhibitory protocols (including low-frequency rTMS and cTBS) demonstrated a *BDNF* polymorphism effect on the response to neuromodulation. A previous systematic review, which only included healthy individuals, found approximately 20-30% of studies showed *BDNF* polymorphism-related changes in MEP^59^. When considering only the studies with healthy individuals in the current systematic review, 12 out of 23 (52.2%) protocols (excitatory: 6; inhibitory: 6) demonstrated *BDNF* polymorphism-related changes, which is considerably more than the previous systematic review^59^. This is possibly due to diverse statistical approaches, as the previous systematic review only considered the results of ANOVA to determine whether there was a *BDNF* polymorphism effect, while the current study included any statistical analyses showing differences in outcome measures between *BDNF* genotypes. Another systematic review and meta-analysis study in healthy individuals indicated that the effects of TBS (including both iTBS and cTBS) favored Val/Val homozygote over Met allele carriers, with the excitatory iTBS showing greater effect sizes^53^. While the current systematic review did not compare the effect sizes across the included studies, we expanded the scope to also include pathological populations beyond healthy individuals. A *BDNF* polymorphism effect was evident in 17 out of 22 (77.3%) protocols in pathological populations, with a marginally higher percentage in the excitatory (10 out of 12, 83.3%) compared to inhibitory (7 out of 10, 70%) protocols.

### 4.4 Methodological Considerations

The methodologies and stimulation parameters used in the included studies were variable, likely due to a lack of consensus on optimal protocols in rTMS and TBS research. While the stimulation intensity, stimulation frequency, and pulse number used for TBS were consistent in most studies (80% AMT, 50 Hz, and 600 pulses, respectively), these three parameters were more variable in rTMS studies. Although the consistency of stimulation parameters reduced variability resulting from the neuromodulation device, of the 26 TBS protocols, only 14 studies (53.8%) showed significant *BDNF* polymorphism effects. Personal factors such as symptom severity and demographics need to be taken into account as possible predictors of rTMS responsiveness.

Some commonly used outcome measures were not completely comparable given the different scales or units (e.g., MEP amplitude measured with the original unit in mV, percentage difference, or log-transformed; Hamilton rating scale for depression in 21 versus 17 items). Some studies segregated participants with Val/Met heterozygotes and Met/Met homozygotes as different genotypes, whereas others grouped them as Met allele carriers since there were often not enough data points with the Met/Met homozygotes. This precludes inferring the association between response to rTMS and Val/Met heterozygotes or Met/Met homozygotes.

As for stimulation sites, there were not enough studies using hotspots other than M1 in studies with healthy individuals, and thus whether brain region influences *BDNF* polymorphism effects remain inconclusive. In stroke, *BDNF* polymorphism effects were more apparent when targeting the ipsilesional M1 with high-frequency rTMS^35,36,42^ compared to targeting the contralesional M1 with low-frequency rTMS^30,34,37^ or ipsilesional iTBS^38^. Facilitating the post-stroke ipsilesional hemisphere or inhibiting the contralesional hemisphere was based on the interhemispheric imbalance model that hypothesizes asymmetric interhemispheric inhibition^60^. While administering low-frequency rTMS at the contralesional M1 and high-frequency rTMS at the ipsilesional M1 have been recommended by the most recent guidelines on the therapeutic use of rTMS^7^, it is possible that Met allele carriers are less responsive to high-frequency rTMS and therefore *BDNF* polymorphism should be considered while determining the rTMS protocol to enhance post-stroke motor recovery.

In the risk of bias assessments, lack of allocation concealment was often seen in the included studies, or there was no description of whether the investigators performing TMS-derived assessments were aware of the rTMS/TBS protocol. Other common biases included no blinding of the participants receiving different protocols of rTMS, and incomplete reports of stimulation parameters and the original values of group means in TMS-derived outcomes. Most of the studies (94.3%) did not report procedures or data in at least one domain of risk of bias assessment. These were rated as unclear risk of bias and further contributed to potential bias while interpreting the findings.

### 4.5 Limitations and Future Directions

Some limitations of this systematic review are noted. First, only two studies examined the *BDNF* C270T polymorphism; therefore, there was not enough data to conclude the relationship between C270T polymorphism and response to neuromodulation. Second, most included studies had small sample sizes of less than 50 participants. It is possible that some studies were underpowered with insignificant differences in outcome measures with classification according to *BDNF* genotypes or assignments to receive different rTMS protocols. Third, while *BDNF* polymorphism as a contributing factor to rTMS modulatory effects was widely studied in healthy individuals, there were insufficient publications in people with medical conditions. Quantitative data analysis was limited by the small number of studies, heterogeneity of patient characteristics, unretrievable data of key parameters, and inconsistency of rTMS protocols. Future studies are warranted to explore interactions between *BDNF* polymorphism and rTMS parameters and dosages while controlling participant-specific factors, such as symptom severity, in populations with medical conditions.

## Data Availability

All data produced in the present study are available upon reasonable request to the authors

## Notes

### Competing Interest Statement

The authors have declared no competing interest.

### Funding Statement

Hendricks Intramural Seed Grant, SUNY Upstate Medical University

### Author Declarations

This systematic review used articles published on existing public datasets.

